# Risk-benefit balance with metformin treatment post pregnancy in breastfeeding women - the transfer of metformin into human breast milk and plasma of the child – a low intervention clinical trial protocol

**DOI:** 10.1101/2025.04.02.25325122

**Authors:** Linda Englund-Ögge, Jarl Hellman, Helena Backman, Jennifer Drevin, Erik Melander, Erica Sundell, Mats Hansson

## Abstract

**Aims:** The primary aim is to determine the concentration of metformin in the plasma of breastfed infants to lactating women being treated for type 2 diabetes.

The secondary aim is to determine the concentration of metformin in breast milk and maternal plasma and the milk-to-plasma ratio of the mothers and to calculate the average daily infant dose (ADID) and relative infant dose (RID).

**Methods:** The study has a low intervention clinical trial design in the sense that breast milk and blood will be collected merely to study excretion of metformin into breastmilk and transferal to her child. Participation in the study will not decide or in any other way interfere with patients’ treatment as prescribed by their physician. Only patients that already have been assigned treatment with metformin by their physician will be approached and asked for participation. All procedures for blood collection will follow established clinical routines at the clinical sites. The sampling will take place ≈ 6-8 weeks postpartum.

The study has been approved as a low-intervention clinical trial by the Swedish Medical Product Agency (Publicly available in CTIS (EU CT no. 2022-501693-19-00 and registered in EUPAS 105190) which includes an approval by the Swedish Ethical Review Authority. The date for the ethics approval is 17 February 2023

**Results:** Up to date (24 February) 18 women and infants have been recruited and sampled. Recruitment is ongoing.

**Conclusions:** An evaluation of pharmacokinetic results as well as clinical experiences related to recruitment and sampling will be systematically conducted upon completion of the study, which is estimated to conclude by the end of 2025. Preliminary experiences suggest that the primary challenge for a study of this nature is recruiting a sufficient number of participants.

**Highlights:** With as few as 5% of available medications being adequately monitored, tested and labelled with safety information for use in breastfeeding women there is a great need to monitor and evaluate the potential risk of transfer of medicines to the infant. We demonstrate here how a low-intervention clinical trial may be designed in order to provide evidence of transfer.

## Introduction

Annually, over 5 million women become pregnant in the European Union (EU), and the majority of them use at least one medication during pregnancy and breast feeding (1). However, only a small portion of available medications have been adequately monitored, tested, and labeled with safety information specific to use during pregnancy and breastfeeding 2,3). In spite of the lack of comprehensive data, these medications are sometimes utilized during pregnancy and lactation when the anticipated benefits are considered to outweigh the potential risks. Depending on medications, the risk situation is complex. There is a risk/benefit balance to be made by the treating doctor related to the medication with acknowledgement of known adverse reactions but there is also a risk associated with the disease itself (see below). This calls for the involvement of clinicians, both from antenatal care units, and specialist care units such as obstetric and neonatology departments, who often meet and treat pregnant or lactating women, and i.e. in the case of diabetes, mostly endocrinologists.

In clinical practice, it is tempting to adopt a precautionary approach by discontinuing the prescription of inadequately tested medications during pregnancy and breastfeeding or by advising mothers to cease breastfeeding. This approach carries, however, significant consequences for both the mother and her child. The mother may have a legitimate medical need for the medication, which may not be easily substituted with an alternative treatment. Furthermore, it is well established that breastfeeding has numerous benefits for both maternal and infant health. The problem at focus here, is that the lack of conclusive evidence regarding the transfer of metformin to breastfeeding infants could result in increased risks for both mothers and their children. Anecdotal evidence from doctors in Sweden indicate that clinical practice differs, some withdraw the drug, others continue.

In order to obtain reliable data on prescriptions, we requested the National Board of Health and Welfare to conduct a cross-match between the National Prescribed Drug Register (NPDR) and the National Medical Birth Register (NMBR) for 45 different medications that, according to the European Network of Teratology Information Services (ENTIS), lack sufficient scientific evidence for use during pregnancy and breastfeeding. A threshold of 500 prescriptions per year for the entire country of Sweden was established, where metformin was one of the 12 medications identified through this process.

### Metformin use during pregnancy and childbirth

Type 2 diabetes (T2D) is a chronic metabolic disease characterized by both insulin resistance and impaired insulin secretion, which leads to elevated blood glucose levels. Diabetes is a common cause of visual impairment and blindness, neuropathy, kidney failure, myocardial infarctions, strokes, as well as diabetic foot ulcers and lower limb amputations. Regular physical activity, a healthy diet, maintaining a normal body weight are ways to prevent or delay the onset of type 2 diabetes. The preferred treatment in T2D is lifestyle intervention together with medication use and regular screening for complications of the disease.

The past and present escalation in diabetes prevalence is alarming and has been rising more rapidly in low- and middle-income countries than in high-income countries with T2D accounting for about 90-95% of all diabetes cases. A recent pooled analysis of 1108 population-representative studies estimated that 14% of adults (13.9% women and 14.3% men) had diabetes, a substantial increase from 630 million 1990 to 828 million people 2022 (4). The largest increases were in low-income and middle-income countries in Asia, north Africa, Latin America, the Caribbean and the Middle East. In most countries, especially in low-income and middle-income countries, medical treatment for diabetes has not increased sufficiently in comparison with the huge rise in prevalence. Although genetic predisposition partly determines the individual risk of developing T2DM during lifetime, a sedentary lifestyle and unhealthy diet are important drivers of the current global epidemic. Early developmental factors such as intrauterine exposures to maternal diabetes and obesity may also have a role in susceptibility to develop T2DM later in life (5). Another important reason for the increased prevalence of diabetes is an aging world population, nearly half of individuals with diabetes are over the age of 65 (6).

In line with the general increase of T2DM in different populations, T2DM and gestational diabetes during pregnancy have increased (7). Treatment during pregnancy include insulin and/or metformin, depending on different national guidelines (e.g. ADA; NICE). Since insulin does not cross the placenta; the decision regarding whether to continue metformin or to start the treatment during pregnancy is done after information and consent. The question if lactation is recommended with *post-partum* use of metformin is a decision balancing benefits for mother and child with possible to date unknown harms for the neonate/child.

Metformin (dimethylbiguanide) is a member of the biguanide class of medications and originally linked to Galega officinalis, a traditional herbal medicine commonly known as goat’s-rue (8). Originally guanidine derivatives were used to treat diabetes already in the 1920s and 1930s but were discontinued due to toxicity. Metformin was later rediscovered due to a possible antiviral effect against influenza and the French physician Jean Sterne (1909-1997) was first to report the use of metformin to treat diabetes in 1957 (9). Its primary mechanism of action involves inhibiting hepatic glucose production but also enhancing insulin sensitivity. The ability of metformin to reduce insulin resistance without a risk of hypoglycaemia and weight gain gradually increased the usage of metformin in Europe, but it was not introduced into the US until 1995.

The UK Prospective Diabetes Study (UKPDS) was a multicenter study that ran for twenty years (1977 to 1997) in the UK and included 4.000 individuals with newly diagnosed type 2 diabetes with a subpopulation of 342 overweight patients allocated to metformin treatment. Metformin was the first glucose-lowering agent ever reported to improve cardiovascular outcomes. The results from this landmark study were presented in 1998 and metformin then became a first-line agent to manage type 2 diabetes worldwide, a cornerstone of treatment of type 2 diabetes and the most prescribed glucose-lowering medicine worldwide (10). Adverse effects secondary to metformin are primarily related to gastrointestinal disturbances, lactic acidosis is a rare but serious side effect of metformin use. In recent years, metformin has been increasingly questioned as the first-line treatment for type 2 diabetes. However, virtually all major randomized controlled trials involving more modern diabetes medications have included metformin treatment as the foundation of the therapy. Despite extensive use and well documented benefits, a notable gap exists in the literature concerning the safety and efficacy of metformin in specific populations, especially in pregnant and breastfeeding women.

During pregnancy, studies have shown no risk for malformations if metformin is used in early pregnancy, but a concern has been raised regarding the effect of foetal growth and possible risk for being born small for gestational age (SGA), that is known to be a risk factor for later morbidity. The studies showing risk for SGA are predominantly in women that have started metformin in early pregnancy; which suggests that either the duration of treatment, underlying pathology or placental changes could be part of this effect (11). The large, randomized MiG Trial (treatment start midst of pregnancy) was a landmark study that investigated the use of metformin compared to insulin for the treatment of gestational diabetes mellitus. The findings indicate that metformin, whether administered alone or in conjunction with supplemental insulin, is an effective and safe treatment option for women with gestational diabetes who meet the standard criteria for initiating insulin therapy (12). Similarly, the extensive international randomized multicenter MiTy Trial (Type 2 diabetes and early newly detected overt diabetes before gestational week 20) observed several benefits related to maternal glycemic control and neonatal adiposity, resulting in fewer large infants but a higher proportion of small-for-gestational-age infants among those treated with metformin (13). The randomized EMERGE trial was the first to administer metformin early in pregnancy at the time of gestational diabetes diagnosis, demonstrating that metformin was safe and associated with a reduction in the incidence of large-for-gestational-age infants but also associated with lower CRL measure in early pregnancy indicating an early effect on fetal size (14). In a prespecified secondary analysis of EMERGE there was no significant increase in the odds of having an SGA infant with metformin (15). Additionally, the randomized MOMPOD Study examined the effects of metformin in pregnant women with type 2 diabetes, including those with preexisting early gestational diabetes. While the addition of metformin to insulin therapy did not reduce the frequency of composite adverse neonatal outcomes, compared to placebo, women randomized to metformin were less likely to deliver large-for-gestational-age infants, with the proportions of small-for-gestational-age infants remaining similar (16).

Although metformin use is considered safe for mothers, concerns persist regarding its long-term metabolic effects on children exposed to the drug in utero, as metformin, unlike insulin, crosses the placenta (17). The use of metformin for the management of gestational diabetes mellitus (GDM) remains a contentious issue, with varying clinical recommendations across different countries. Metformin is endorsed by the Swedish National Board of Health for managing gestational diabetes mellitus (GDM) and type 2 diabetes during pregnancy (18). Metformin is also a commonly used medication in polycystic ovary syndrome (PCOS).

### Metformin transfer - what is already known?

Metformin crosses the placenta but there is limited data concerning the transfer of metformin to human breast milk and its effects on breastfeeding infants. We identified five clinical publications covering six different studies (18-22) that have investigated the excretion of metformin in maternal blood and breast milk, as well as health-related outcomes for breastfeeding children. Five studies have estimated metformin concentrations in plasma or serum after recruiting between three to seven lactating women each (18,20,21). The participants had been treated with metformin for pre-existing type 2 diabetes, gestational diabetes mellitus, or PCOS and had reached a steady state concentration of metformin, with exception from one study (study II, Gardiner et al, (20). The metformin dosages administered ranged from 500 mg to 3000 mg per day, with a few participants also using extended-release formulations. The lowest reported maternal peak plasma concentrations were 0.5-0.7 mg/L (13), while the highest reached 1.61 mg/L (20). The peak concentrations in plasma or serum occurred between 1.8 and 4 hours post-ingestion, with reported half-lives in serum/plasma ranging from 2 to 6.3 hours (18,20,22).

Breast milk sampling taking place 4 days to 15 months postpartum show mean or median concentrations of metformin ranged between 0.17 mg/L and 0.42 mg/L (18,20,22). Concentration graphs in breast milk exhibited a relatively flat curve, noting a mean difference of 21% between through and peak concentrations in one of these studies (16). The estimated half-life of metformin in breast milk is approximately 14-15 hours (study II, Gardiner et al, 19). The reported milk-to-plasma area under the curve ratios varied among participants, ranging from 0.27 to 0.71, with study mean values between 0.37 and 0.54 (18,20,22). These six different studies have documented health data regarding breastfeeding infants of mothers using metformin, the children involved ranged in age from 4 days to 15 months and weighed between 2.8 kg and 15 kg. Plasma concentrations of metformin were measured for six infants in total, of which metformin was not detected in four of them (20,21). The two infants with detectable metformin levels exhibited plasma concentrations of 0.08 mg/L and 0.05 mg/L, measured 5.3 and 6 hours, respectively, after maternal dosing (18). Estimated daily doses for the infants ranged from 0.014 mg/kg/day to 0.070 mg/kg/day, with mean values between 0.030 and 0.040 mg/kg/day (18,20). The mean relative infant doses, normalized to maternal weight, have ranged from 0,18% to 0,65% (12-14,17). Additionally, Briggs et al. (20) measured blood glucose levels in three breastfeeding infants, finding them to be within normal ranges (47-77 mg/dL). Breastfeeding infants whose mothers used metformin (n=61) demonstrated a normal development, showing no significant differences in length, weight, or motor-social development during their first 18 months compared to non-breastfed children. No evidence of adverse effects among the participating children were reported.

Thus, until today, only a limited number of publications have investigated the effects of maternal metformin treatment on breastfeeding and the health of breastfeeding children. To thoroughly assess any potential risks to breastfeeding infants, it is essential to study the excretion of metformin in breast milk and to also measure the concentrations of metformin in the plasma of breastfeeding infants. In summary, there is a need for more conclusive evidence regarding the transfer of metformin to infants via breast milk.

### Setting up a low-intervention clinical trial

In order to assess the transfer of metformin to the infant of breastfeeding mothers we set up a mother-infant pair lactation study with sampling of breast milk, mothers’ plasma and infants’ plasma. The study has a low intervention trial design in the sense that breast milk and plasma are collected merely to study excretion of metformin into breastmilk and transferal to the child. Participation in the study does not decide or in any other way interfere with the patients’ treatment as prescribed by their physician. Only patients that already have been assigned treatment with metformin by their physician are approached and asked for participation.

Clinical praxis is followed in all procedures, but since plasma samples of newborns are not taken on a routine basis at the desired period of 6-8 weeks postpartum a low-intervention clinical trial application was warranted.

The clinical trial is approved by the Swedish Medical Product Agency (Dnr. 5.1.1-2023-0905929). The clinical trial protocol is available via the Clinical Trial Information System at the European Medicine Agency (No. 2022-501693-19-00). It is also registered and publicly accessible at the EU PAS Register (EUPAS 105190).

Taking plasma samples from newborn infants requires close collaboration with experienced and dedicated clinical centers. Uppsala University is the sponsor of the study, with funding via the IMI-ConcePTION project (Innovative Medicines Initiative 2 Joint Undertaking, Grant agreement No. 821520). The trial is designed in compliance with the EU regulation on low-intervention clinical trials on medicinal products for human use (536/2014), the Declaration of Helsinki, ICH-GCP standards applicable to the trial procedures, and current national regulations governing this clinical trial.

### The low-intervention trial

Real world data about transfer of medicine to an infant in association with breast feeding requires plasma sampling from the infant. We present how the study is designed, the methodology, sampling procedures, informed consent procedures, data handling and ethical requirements managed. The samples are collected and biobanked for pharmacokinetic analyses and, following approved information and consent procedures, to be used for future research as approved by appropriate ethical review boards. Sampling is still ongoing.

According to FDA recommendations for clinical lactation standards, there are several study designs that may be used according to ethical requirements related to the burden of data collection on the mother while still obtaining adequate data. Access to study population, half-live of the drug, stability of the drug during collection procedures and possibility to provide pharmacokinetic data are other criteria for selecting an appropriate study design (24). Different designs (milk-only, mothers milk and plasma, mother-infant pair) are possible and permitted.

This project aims to demonstrate the proper methodologies for biobanking and the sampling of breast milk and blood plasma to ensure that the resulting analytical data comply with regulatory requirements from the EMA and FDA respectively. This initiative holds significant potential for various contexts where there is currently insufficient evidence regarding the transfer of medications to the child during breastfeeding, a concern that applies to nearly all medications. By establishing a biobank for breast milk and associated samples of blood plasma samples for research purposes, there is also an opportunity to investigate in the future how e.g., genetic factors may influence drug transfer. Furthermore, this biobank could facilitate the future collection of data to monitor the effects on the children.

#### Benefit-Risk Evaluation

The project will not alter the woman’s treatment in any way; thus, participation will not expose her to any additional risks. Thus, there is no change regarding the prescription of metformin, but this is done according to the physician’s regular prescription. The risk-benefit balance associated with metformin is not altered by participation in this project. Blood sampling will be conducted according to established clinical routines. Participants may experience a slight sting during the procedure, and there is a potential for bruising. Additionally, there is a minimal risk of nerve damage, which could manifest as pain or numbness. When blood sampling involves children, there are particular concerns related to pain. The clinically validated protocol utilized in this project involves obtaining a venous blood sample (0.8 ml) from the back of the hand. To alleviate discomfort, lidocaine/prilocaine cream (EMLA®) and/or a sugar solution is administered (25). This procedure is performed by experienced healthcare professionals. A total of 28 ml of maternal blood is collected, divided over two occasions, which is significantly less than the 450 ml typically drawn during a standard blood donation.

Only data relating to the blood sampling procedure and information concerning any additional medications the woman is currently taking is collected. The integrity risk is minimized by pseudonymizing the data and storing the data and the coding key separately. Research participants are not anticipated to receive any direct benefits from their involvement in the study.

The study results will be able to contribute information that in the future can help breastfeeding women to make an informed decision about using metformin in connection with breastfeeding. Such information will also help healthcare professionals to provide evidence-based advice to breastfeeding women about the use of metformin. An alternative to obtain data regarding the transfer of drugs during breastfeeding is to only collect breast milk with estimations about drug transfer to the infant based on pharmacokinetic modeling. However, it is only when you also take a blood sample from the breastfed child that you get reliable data on how much of the medicine is actually transferred to the child. We suggest that this move from estimation to measurement as the “golden standard” that should apply to lactation studies internationally.

#### Trial design and procedures

The trial is a low intervention trial with biobanking of breast milk and plasma. Collection of samples is made at two clinical sites, the Specialist-antenatal Breast Department of Obstetrics, Sahlgrenska University Hospital, Östra, in Göteborg and the Breast-feeding Center, Department of Obstetrics, Örebro University Hospital.

General information about the project is provided in Swedish, Arabic, Persian, Somali and English through a Facebook group of patients with diabetes, hosted by the Swedish patient organisation *Diabetesf*ö*rbundet*. Additionally, information is distributed through a network of healthcare professionals via *Janusmed* at Region Stockholm, to Swedish physicians specializing in diabetes care, and through social media channels, posters displayed at obstetric clinics, maternity wards, and endocrinology clinics throughout Sweden. A video is also distributed via these platforms, along with a link to the project webpage for further information. Specific oral and written information about the study is provided through obstetric departments and affiliated maternity clinics, childcare centers, and breastfeeding support centers. Breastfeeding women treated with metformin for suspected or diagnosed type 2 diabetes mellitus (T2DM), of 18 years of age and older with infants are invited to participate in the study.

The principal investigator at each site will ensure that participants receive comprehensive oral and written information about the trial, including its purpose, associated risks and benefits, and the criteria for inclusion and exclusion. Participants are informed that they have the right to withdraw from the trial at any time without needing to provide a reason. They will also have the opportunity to ask questions and be given adequate time to consider the information presented. If they choose to participate, individuals will be required to sign an informed consent form. Additionally, participating women will be asked to have their partners (other caregivers) sign the consent form. The signed form will be brought to the medical doctor at the sampling center who will sign the form. A copy of the participant information as well as the informed consent form will be provided to the woman. Each participant will be assigned a unique subject number, which will be recorded on a subject identification list.

All blood collection procedures follow established clinical routines at the participating clinical sites. The sampling takes place once at about 6-8 weeks postpartum. Breast milk is collected twice during this visit using an electric breast pump. Venous blood samples are collected from both the mother and the infant. After centrifugation, plasma and breast milk samples are aliquotated, frozen at −80 degrees Celcius, and stored at Biobank Väst and Örebro Biobank, respectively. Subsequently, they are transferred to Uppsala Biobank. The samples are analyzed for pharmacokinetic properties using mass spectrometry at the UDOPP Platform (Uppsala Drug Optimization and Pharmaceutical Profiling) within the Department of Pharmacy at Uppsala University.

#### Overview of sampling procedures

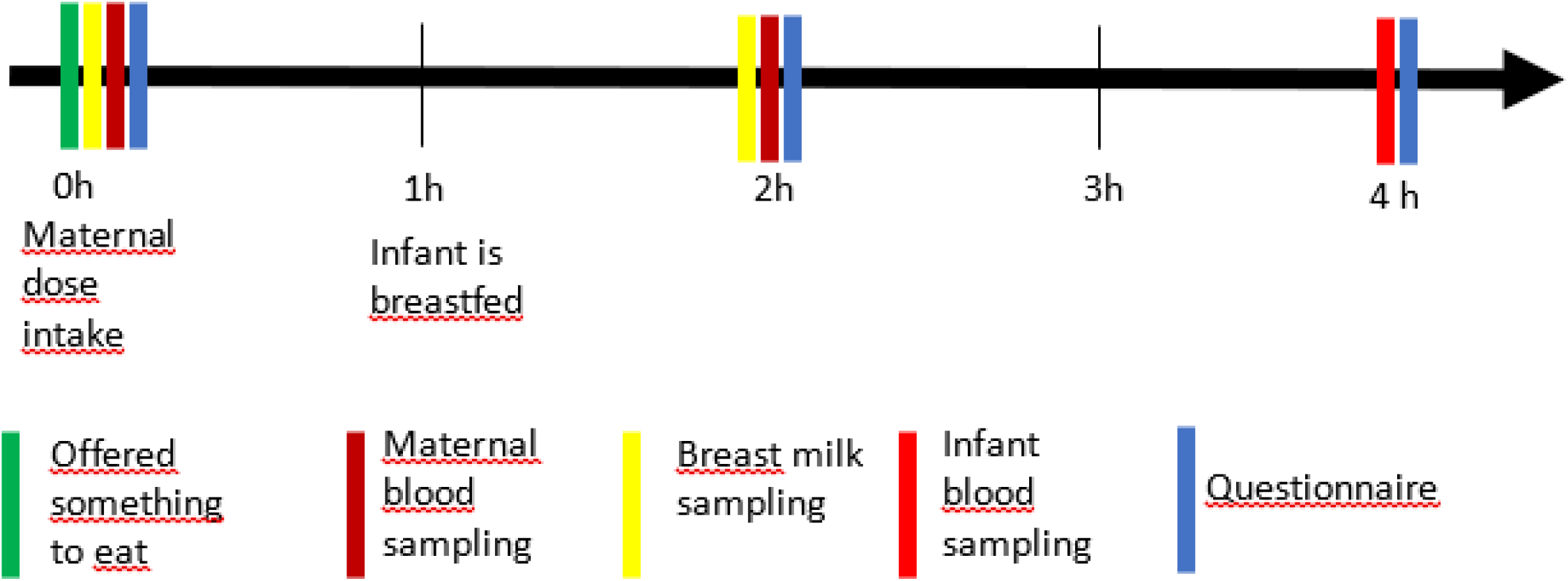

#### Measurements

Regarding pharmacological safety, the project will measure pharmacokinetic data of breast milk and blood related to a medication (metformin) that is already approved and administered according to standard clinical procedures for women outside the protocol of this project. Adverse events that may impact on breastfeeding and/or breastmilk, e.g., mastitis, will be documented. The maternal dosage will be calculated in mg/kg/day. The Absolute Infant Dose (AID) in mg/kg/day will be estimated using a standard daily milk intake of 200 mL/kg/day, in accordance with FDA guidelines for lactation studies (24). While an estimated milk intake of 150 mL/kg/day is a reasonable assumption for calculating daily infant dosage, greater volumes are common in early infancy and often correlate with the period during which most reported infant adverse drug events occur. Therefore, estimates of infant risk should also consider a milk intake of 200 mL/kg/day in early infancy. The Relative Infant Dose (RID) and the maternal metformin milk-to-plasma ratio will also be calculated, and metformin concentration in the infant’s blood will be reported.

#### Data protection

Subjects are informed about how their trial data will be collected, used and disclosed. The content of the informed consent form complies with relevant privacy and data protection legislation, including the Regulation (EU) 2016/679 (General Data Protection Regulation, GDPR). The subject information and the informed consent form explain how trial data are stored to maintain confidentiality in accordance with national data legislation and GDPR (see section 14). All information processed by the sponsor is pseudonymized and coded.

Sampling details and health-related details are collected using a paper questionnaire and then registered in the program REDCap (Research Electronic Data Capture). Informed consent forms are sent by registered mail to Uppsala University and stored in a locked cabinet only accessible by authorized personnel. Data are uploaded in an encrypted format and hosted on a secure server at Uppsala University (Dataportal ALLVIS) and Region Uppsala, in accordance with an established process approved by the Swedish Ethical Review Authority.

### Preliminary findings

Up to date (24 February 2025) 15 women and infants have been recruited and sampled in Göteborg. 3 women and their infants in Örebro. These samples have been transferred to Uppsala Biobank and delivered to UDOPP for analysis. Recruitment is ongoing at both sites.

An evaluation of clinical experiences related to recruitment and sampling will be systematically conducted upon completion of the study, which is estimated to conclude by the end of 2025. Preliminary experiences suggest that the primary challenge for a study of this nature is recruiting a sufficient number of participants. The underlying reasons for this difficulty are, as described above, that a substantial number of women have other newer medications for type 2 diabetes, metformin could be discontinued by prescribing physicians, or women do not want to or are advised against breastfeeding. After *partus* and during breastfeeding, insulin sensitivity is often increased, which may temporarily reduce the need for diabetes medication. Concurrently, the prevalence of type 2 diabetes mellitus (T2DM) is on the rise. A significant insight from this study, besides evidence on metformin transfer, could be the necessity of involving more clinical centers in the recruitment process.

## Data Availability

No datasets were generated or analysed during the current study. All relevant data from this study will be made available upon study completion.

## Acknowledgements

This study is a part of IMI-ConcePTION. The ConcePTION project has received funding from the Innovative Medicines Initiative 2 Joint Undertaking under grant agreement No. 821520. This Joint Undertaking receives support from the European Union’s Horizon 2020 research and innovation program and EFPIA.

This study is conducted using professional biobank services from Biobank Väst, University of Gothenburg, Core Facilities at Sahlgrenska Academy, Örebro Biobank, Uppsala Biobank and Biobank Sweden.

---

**Fig 1.**
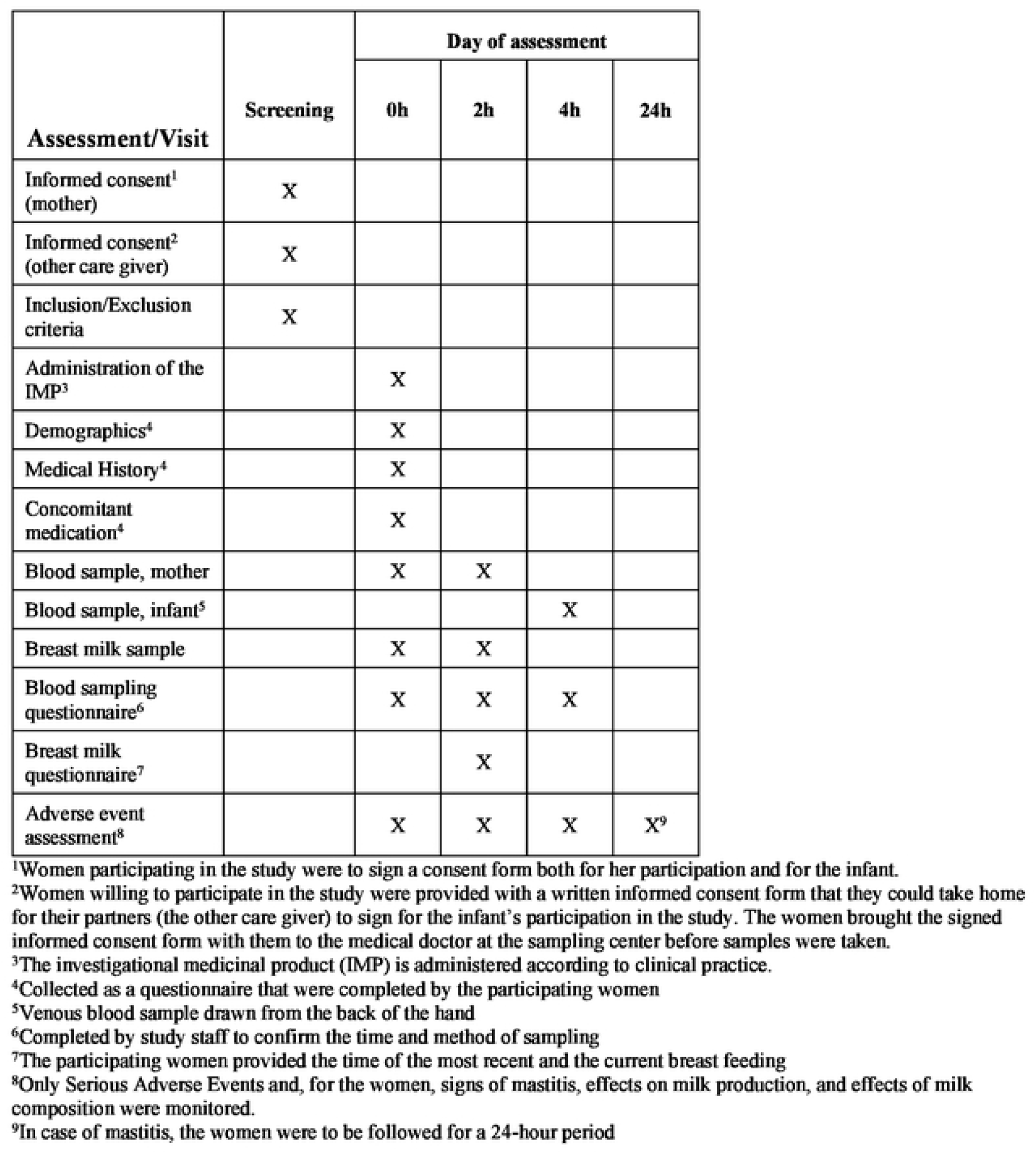
SPIRIT Schedule.

